# Emergence and spread of the potential variant of interest (VOI) B.1.1.519 predominantly present in Mexico

**DOI:** 10.1101/2021.05.18.21255620

**Authors:** Abril Paulina Rodríguez-Maldonado, Joel Armando Vázquez-Pérez, Alberto Cedro-Tanda, Blanca Taboada, Celia Boukadida, Claudia Wong-Arámbula, Tatiana Ernestina Nuñez-García, Natividad Cruz-Ortiz, Gisela Barrera-Badillo, Lucía Hernández-Rivas, Irma López-Martínez, Alfredo Mendoza-Vargas, Juan Pablo Reyes-Grajeda, Nicolas Alcaraz, Fernando Peñaloza-Figueroa, Dulibeth Gonzalez-Barrera, Daniel Rangel-DeLeon, Luis Alonso Herrera-Montalvo, Fidencio Mejía-Nepomuceno, Alejandra Hernández-Terán, Mario Mújica-Sánchez, Eduardo Becerril-Vargas, José Arturo Martínez-Orozco, Rogelio Pérez-Padilla, Jorge Salas-Hernández, Alejandro Sanchez-Flores, Pavel Isa, Margarita Matías-Florentino, Santiago Ávila-Ríos, José Esteban Muñoz-Medina, Concepción Grajales-Muñiz, Angel Gustavo Salas-Lais, Andrea Santos Coy-Arechavaleta, Alfredo Hidalgo-Miranda, Carlos F. Arias, José Ernesto Ramírez-González

## Abstract

SARS-CoV-2 variants have emerged in late 2020 and there are at least three variants of concern (B.1.1.7, B.1.351, P1) reported by WHO. These variants have several substitutions in the Spike protein that affect receptor binding; they present increased transmissibility and may be associated with reduced vaccine effectiveness. In the present work, we are reporting the identification of a potential variant of interest harboring the mutations T478K, P681H, and T732A in the Spike protein, within the newly named lineage B.1.1.519, which rapidly outcompeted the preexisting variants in Mexico and has been the dominant virus in the country during the first trimester of 2021.

---

Viral mutation is a natural and expected event generated during genomic replication and interaction with the host, resulting in the occurrence of genetic groups also called lineages. The latter differ from each other by specific mutations that accumulate over time, causing the appearance of variants. A variant could be defined as a virus with specific genetic mutations that differ from the original virus, reflecting in some cases SARS-CoV-2 adaptation to its novel human host. Although the majority of mutations in the SARS-CoV-2 genome are expected to be neutral or deleterious, some mutations can confer a selective advantage and may be associated with enhanced fitness, increased infectivity and/or immune evasion [1, 2, 3]. Importantly, the emergence and spread of variants associated with changes in transmission, virulence and/or antigenicity can impact the evolution of the COVID-19 pandemic and might require appropriate public health actions and surveillance [4].

New SARS-CoV-2 variants are spreading rapidly around the world, becoming a public health concern. As of February 23, 2021, Pan-American Health Organization (PAHO)/World Health Organization (WHO) and Global initiative on sharing all influenza data (GISAID) reported the appearance of at least three Variants of Concern (VOC) that have presented characteristics with implication in public health. Variant B.1.1.7 was identified for the first time in the United Kingdom in September 2020 [4, 5], and by December 2020 it represented 43% of the genomes sequenced, increasing to 82% in January 2021 and to 94% in February 2021 [6]. This variant is of growing concern since it has been shown to be significantly more transmissible than other variants [7], and to likely have increased severity, based on hospitalization and fatality rates. Variant B.1.351 was detected for the first time in South Africa, in 64% (261 of 411 genomes) of the sequences reported in December 2020, increasing to 75% (99 of 132 genomes) in the next month [6]. Epidemiological data analysis estimated that this VOC is 50% more transmissible than the previous circulating variants. Finally, the variant was detected for the first time in Brazil in 47% (61 of 130) of the viral genomes in December 2020, increasing to 74% (111 of 150 genomes) in the next month [6, 8]. Of relevance, it has shown a reduced neutralization by convalescent and post-vaccination sera. These SARS-CoV-2 VOCs have independently acquired some of the same Spike protein mutations, particularly E484K, N501Y, S477N, and K417T, which have been associated with increased viral transmission and / or decreased sensitivity to antibody neutralization [9].

In Latin America, with the exception of P.1 and P.2 observed in Brazil, no other variants with the potential of rapid expansion have been reported so far [10]. Here we report the identification of a potential VOI harboring the mutations T478K, P681H, and T732A in the Spike protein, within the newly named lineage B.1.1.519, derived from the B.1.1.222 lineage, which rapidly outcompeted the preexisting variants in Mexico and has been the dominant virus in the country during 2021.

Derived from the genomic surveillance carried out in Mexico, 2,692 genomic sequences were obtained in this study and are part of the 3,156 sequences deposited in the GISAID from March 1, 2020 to March 21, 2021. As a result of the analysis of this set of sequences, we observed the presence of 91 PANGO lineages, being B.1.1.519 (37.8%), B.1 (13.9%), B.1.1.222 (10.3%), B.1.1 (5.7%), B.1.609 (5.6%) and B.1.243 (4.5 %) the most prevalent.

A striking observation was the detection of the B.1.1.519 lineage, derived from B.1.1.222, that harbors a mutation T478K in the Spike protein. This variant had not been detected in Mexico before October 2020, when it was found in Mexico City, and phylogeographic analyses suggest that the B.1.1.519 variant emerged around mid-September 2020 (9). In November 2020, 13% (16/123) of the characterized cases of COVID-19 were caused by this variant, and in December this proportion increased to 29.3% (97/331). In January 2021, the percentage of B.1.1.519 rose to 51.5% (229/445), increasing in incidence to 73.6% (808/1098) in February. On the other hand, a decreasing frequency of the B.1 lineage that had predominated in Mexico in 2020 was observed, going from 36.27% (284/783) between March and September, to 2.37% (26/1098) in February 2021.

Since the identification of B.1.1.519 in October 2020, a total of 1,195 genomic sequences have represented this variant, of which the majority are from Mexico City and are spread in other 24 states of the Mexico (Aguascalientes, Baja California, Baja California Sur, Campeche, Chiapas, Chihuahua, Coahuila, Colima, Durango, Guerrero, Hidalgo, Jalisco, Morelos, Nuevo León, Oaxaca, Puebla, Querétaro, Quintana Roo, San Luis Potosí, Estado de Mexico, Tamaulipas, Tlaxcala, Veracruz, Yucatán, and Zacatecas).

A detailed analysis of the samples from Mexico City indicates that in November B.1.1.519 was present in 17.8% (13/73) of the cases, while in December 2020 this proportion increased to 47.5% (47/99). In January 2021, the variant was detected in 77.5% (138/178) of the cases and by February in 90.9% (349/384). This significant increase in the frequency of B.1.1.519 in Mexico City outcompeted preexisting variants between October 2020 and February 2021, as shown in Figure 1. This increase was also observed in other regions of the country (Figure 1), representing more than 50% of the characterized viruses in some states during the first trimester of 2021. In particular, the variant was highly prevalent in Baja California Sur (51.3%, 20/39), Guerrero (70%, 21/30), Hidalgo (72.2%, 13/18), Morelos (67.3%, 33/49), State of Mexico (83.5%, 76/91), Oaxaca (51.3%, 19/37), Puebla (78.5%, 77/98), Queretaro (70.8%, 34/48), San Luis Potosi (70%, 35/50), and Veracruz (69.5%, 80/115).

**Figure 1.**
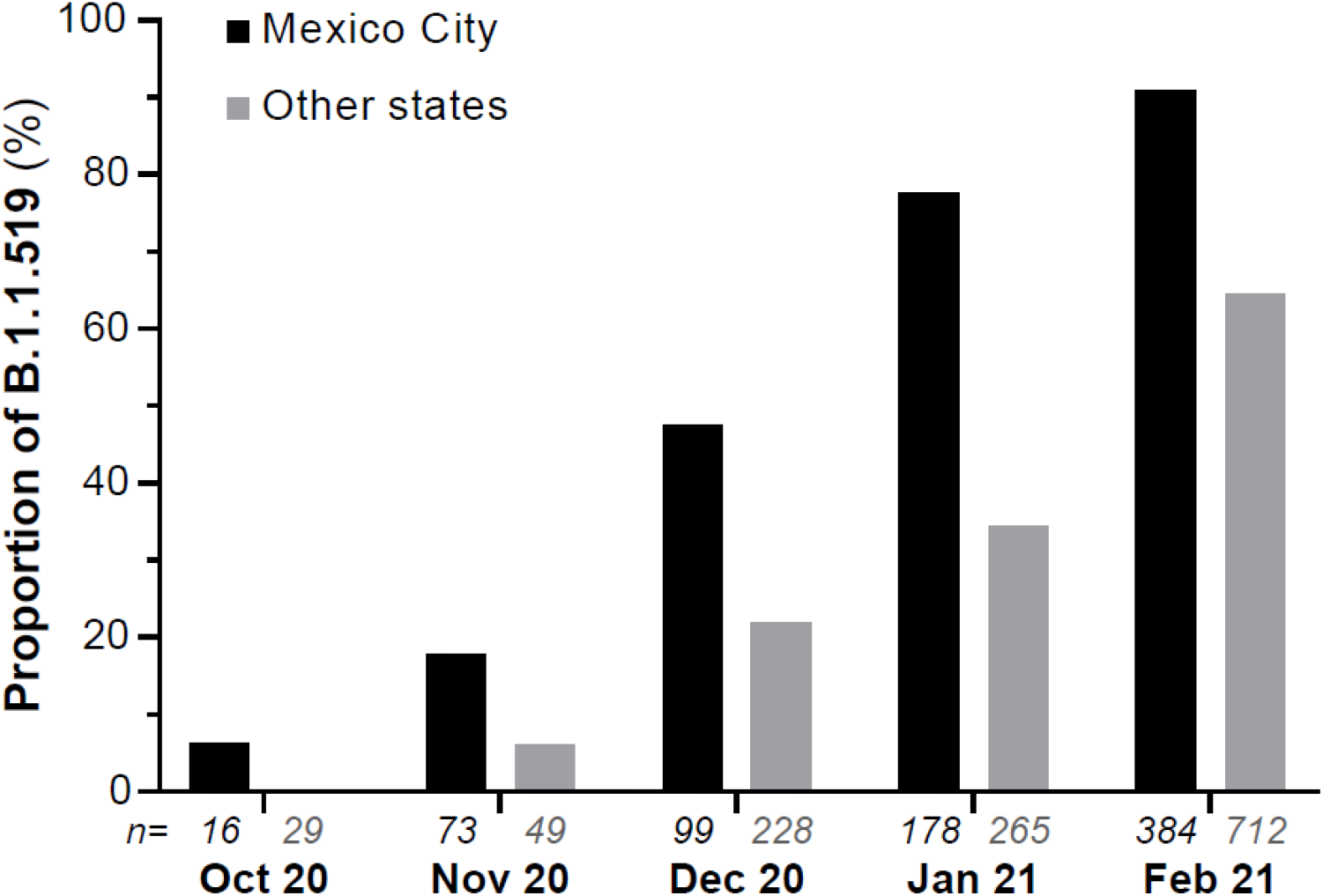
Relative frequency of variant B.1.1.519 from October 2020 to February 2021. The monthly numbers of complete genome sequences (n) obtained from samples collected in Mexico City and other states of Mexico are indicated below each bar.

This variant has also been detected in 17 countries from all five continents. In the Americas, it has been reported in Canada, Brazil and the USA, and recently it was detected in Brazil, Chile, Aruba, Martinique and Curazao [11]. However, currently this variant is not predominant in these countries.

The overall genome analysis of the viruses in the B.1.1.519 lineage showed the presence of 20 mutations in total, compared to the Wuhan-Hu-1 reference genome (NCBI accession number MN908947). Eleven of these mutations are non-synonymous and four of them are present in the Spike protein. Notably, a T478K mutation is present in its receptor binding domain (RBD), where mutations have been shown to reduce the activity of some monoclonal antibodies (9). All amino acid and nucleotide changes are listed in Table 1.

**Table1.** Amino acid and nucleotide changes. Main amino acid substitutions are marked in bold

| Mutations |  |  |
| --- | --- | --- |
| Nucleotide | Amino acid | Gen |
| C203T<br>C222T<br>C241T | --<br>--<br>-- |  |
| C3037T<br>C3140T<br>C10029T<br>C10954T<br>A11117G<br>C12789T | --<br>P141S<br>T492I<br>--<br>I49V | ORF1a |
| C14408T<br>T19839C<br>C21306T | P323L<br>--<br>-- | ORF1b |
| C22995A<br>A23403G<br>C23604A<br>A23756G | **T478K**<br>D614G<br>**P681H**<br>**T732A** | Spike |
| G28881A<br>G28882A<br>G28883C<br>C29197T | --<br>R203K<br>G204R<br>-- | N |

The current B.1.1.519 lineage, presented by the vast majority of the reported Mexican sequences, was first identified as a B.1.1.222 lineage. However, harboring mutations T478K, P681H, and T732A clearly differentiated it from this lineage, which does not contain these mutations, giving origin to the B.1.1.519 one. A phylogenomic analysis of genomic sequences using the Nextstrain tool, showed that viruses in the lineage B.1.1.519 (B.1.1.1.222+T478K+P681H+T732A) group independently of the lineage B.1.1.222 sequences, strongly suggesting that this variant should be classified as a Variant of Interest (VOI) (Figure 2). On the other hand, viruses in the lineage B.1.1.519 are already grouped by the GISAID platform in an independent clade, invariably harboring the three mutations mentioned above.

**Figure 2.**
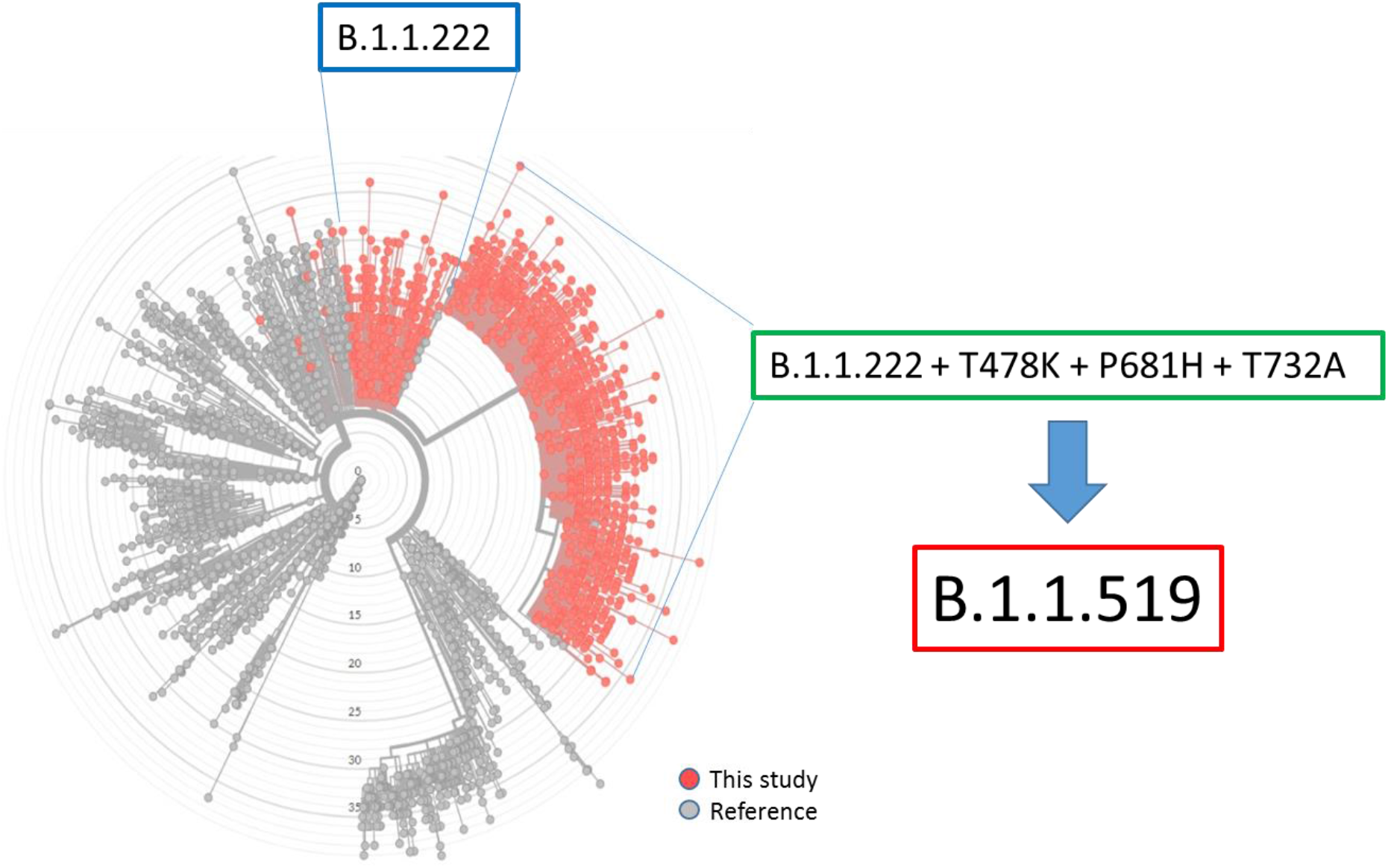
Phylogenomic analysis of SARS CoV-2 sequences obtained in this study (red dots) and of reference sequences (gray dots), performing the clustering of variant B.1.1.519 (red box) independently of the lineage B.1.1.222 (blue box). Viruses in the B.1.1.519 lineage were initially classified within the B.1.1.222 lineage harboring the mutations T478K, P681H and T732A (green box). Phylogenomic tree was powered CC-BY-4.0 license and attribution of nextstrain.org.

An “*in silico*” analysis using different potent structures of related strains, suggested that the position of the T478K mutation in the “S” protein is involved in antibody recognition and the receptor binding site [12]. In a deep mutational scanning of the SARS-CoV-2 receptor binding domain, the T478K mutation did not have a significant effect on folding and human angiotensin-converting enzyme 2 (ACE2) binding [13]. However, this mutation may be involved in immune evasion, particularly escape from antibody neutralization [14]. The P681H mutation is one of the mutations found in the B.1.1.7 variant detected in the UK. According to the definitions described in the document issued by the WHO “Covid-19 Weekly Epidemiological Update” of February 25, 2021, with the special edition of “Proposed working definitions of SARS-CoV-2 Variants of Interest and Variants of Concern”, we can consider the lineage B.1.1.519 potentially as a variant of Interest [15].

Finally, two variants with interesting features were identified in this study. First, 13 sequences belonging to the B.1.1.222 lineage without the T478K mutation, but harboring the T732A mutation and the 69-70 deletion in the Spike protein, the latter being a characteristic mutation of the B.1.1.7 VOC first detected in the UK. In the second instance, 11 sequences corresponding to four lineages different to B.1.1.519 (B.1, B.1.1.222, B.1.1.322 y B.1.323), but containing the same T478K, P681H and T732A mutations in the spike glycoprotein as the variant B.1.1.519. Keeping track of the incidence of these two variants is considered during genomic surveillance.

So far, we do not have experimental evidence to determine if the mutations described here could be associated with changes in transmission, virulence and/or antigenicity or if they could have an impact on the severity of disease, reinfection rates or vaccine effectiveness. For this reason, the importance of a genomic surveillance system, epidemiological studies and experiments to assess viral neutralization of viruses in lineage B.1.1.519 or any new variant, are crucial to investigate the possible biological impact of the mutations in the context of public health.

## Data Availability

All data is available on GISAID website

https://www.gisaid.org/

## Acknowledgements

Authors express their gratitude to Vanessa Rivero, Ariadna Medina, Joaquin Quiroz, Sergio Rangel, Mayra Jiménez, David Fragoso, Nancy Miñoz, Edgar Mendieta, Fabiola Garcés and Adnan Araiza from InDRE for sequencing and bionformatical approach; to Ricardo Grande, Francisco Pulido and Gloria Vazquez from the “Unidad de Secuenciación Masiva y Bioinformática” of the “Laboratorio Nacional de Apoyo Tecnológico a las Ciencias Genómicas” (CONACyT no. 260481) for their support in sequencing services and aslo to working team from molecular biology of the Central Laboratory of Epidemiology IMSS, for technical assistance. We thank to all the staff of the Technological Development and Molecular Research Unit, of the Virology Department and of the Sample Control and Services Department at InDRE for technical assistance.

The findings and conclusions in this report are those of the authors and do not necessarily represent the official opinion of the Ministry of Health in Mexico

## Compliance with ethical standards

### Funding

This work was partially supported by grants “Epidemiología Genómica de los Virus SARS-CoV-2 Circulantes en México” and “Caracterización de la diversidad viral y bacteriana” from the National Council for Science and Technology (CONACyT) of Mexico to C.F.A. and J.A.V.P. respectively and also grants 057, 047 and 072 from the Ministry of Education, Science, Technology and Innovation (SECTEI) of Mexico City to C.F.A., L.H.M. y A.H.M. respectively

### Ethical approval

This article does not contain any studies with human participants or animals performed by any of the authors

